# Impairment of CSF Egress through the Cribriform Plate plays an Apical role in Alzheimer’s disease Etiology

**DOI:** 10.1101/2021.10.04.21264049

**Authors:** Ricardo Zaragoza, Daniel Miulli, Samir Kashyap, Tyler A Carson, Andre Obenaus, Javed Siddiqi, Douglas W Ethell

**Affiliations:** Leucadia Therapeutics Inc; Riverside, CA 92506; Dept of Neurosurgery, Arrowhead Regional Medical Center, Colton, CA; Dept of Pediatrics, University of California Irvine, Irvine, CA; Dept of Surgery, Desert Regional Medical Center, Palm Springs, CA; Developmental Immunology, La Jolla Immunology Institute, San Diego, CA

## Abstract

Cerebrospinal fluid (CSF) clears the brain’s interstitial spaces, and disruptions in CSF flow or egress impact homeostasis, contributing to various neurological conditions. Here, we recast the human cribriform plate from innocuous bony structure to complex regulator of CSF egress with an apical role in Alzheimer’s disease etiology. It includes the pathological evaluation of 70 post-mortem samples using high-resolution contrast-enhanced micro-CT and cutting-edge machine learning, a novel ferret model of neurodegeneration, and a clinical study with 560 volunteers, to provide conclusive evidence of a relationship between cribriform plate aging/pathology and cognitive impairment. Interstitial spaces within the medial temporal lobe and basal forebrain are cleared by CSF flow that drains through olfactory structures to the olfactory bulb—directly above the cribriform plate. We characterized CSF flow channels from subarachnoid spaces under the olfactory bulb to the nasal mucosa through subarachnoid evaginations that subdivide into tiny tubules that connect to an elaborate conduit system within the cribriform plate. These conduits form an internal watershed that runs from the crista galli’s vault to a bony manifold within the olfactory fossa’s back wall, connecting with large apertures in between. We found that the cross-sectional area of apertures limits CSF flux through the cribriform plate, which declines with increasing age. Subjects with a confirmed post-mortem diagnosis of Alzheimer’s disease had the smallest CSF flux capacity, which reduces CSF-mediated clearance in upstream areas and leads to the accumulation of toxic macromolecules that seen AD pathology. We surgically occluded apertures in adult ferrets and found that this manipulation induced progressive deficits in spatiotemporal memory and significant atrophy of the temporal lobe, olfactory bulbs, and lateral olfactory stria. Finally, we explored human cribriform plate aging/pathology and cognition in a clinical study with 560 participants (20-95y). We evaluated cribriform plate morphology with CT and Deep Learning, assessed memory with a novel touch screen platform, tested olfactory discrimination, and asked questions about family history and relevant life events, like broken noses. Deep learning algorithms effectively parsed subjects and established the feasibility of predicting Alzheimer’s disease years before a clinical presentation of cognitive impairment. This study provides a new perspective of the cribriform plate as a critical regulator of CSF egress and brain homeostasis.

## Main Text

Alois Alzheimer first described neuritic plaques and neurofibrillary tangles as defining features of the disease that bears his name in 1907 (Alzheimer, 1907). Yet, more than a century later, it remains unclear whether plaques and tangles are causes or effects of the condition underlying Alzheimer’s disease (AD). Insoluble deposits of amyloid-beta (Aβ) in the dense cores of neuritic plaques have been a significant focus of AD research for over 30 years (Glenner and Wong, 1984; Masters et al., 1984; Tanzi et al., 1987; Selkoe and Hardy, 2016). Mice that over-express high levels of human Aβ develop plaques and display wide-ranging cognitive impairments, but without neurofibrillary tangles or neurodegeneration (Bryan et al., 2009). However, drugs that reduce plaque burden have little impact on cognitive decline (Aisen et al., 2020). An alternative hypothesis suggests that Aβ deposits in the cerebral cortex result from a fundamental biological process gone awry (Ethell, 2014). Therapies that reduce amyloid burden without addressing the underlying process allow pathological progression to continue, with or without plaques, as occurred in anti-Aβ clinical trials (Aisen et al., 2020). A pertinent question is: What biological process(es) could, if disrupted, trigger the hallmarks of AD pathology, including Aβ accumulation, tau hyper-phosphorylation, and neurodegeneration?

Amyloid folds are a structural feature of some proteins that causes them to aggregate at high concentrations, as occurs in amyloidosis (Merlini and Bellotti, 2003). Amyloidogenic proteins accumulate in joints and connective tissues with deficiencies in the clearance of the interstitial compartment. In most of the body, interstitial fluid (ISF) originates as blood plasma from capillaries, except for the CNS, where the blood-brain barrier (BBB) prevents plasma extravasation. Instead, the brain produces cerebrospinal fluid (CSF) as an ISF to clear brain parenchyma. As an incompressible fluid, CSF flow is determined by friction and pressure, which change with movement, position, and cardiac output. Amplified MRI imaging has elegantly shown how the cardiac cycle facilitates CSF movement through the CNS (Terem et al., 2018).

Hydrophobic myelin provides little friction for CSF to flow along white matter tracts compared to hydrophilic gray matter. CSF emerges from the walls of lateral ventricles in the cerebral cortex and flows along white matter tracts, as hydrophobic myelin presents relatively little friction. As CSF reaches cortical gray, its velocity decreases as it passes through narrow interstitial spaces containing proteoglycans. CSF drains into subarachnoid spaces (SAS) at the outermost margin of the brain, where flow velocity can increase. Along the way, CSF accumulates exosomes, debris, and insoluble metabolites that do not efficiently cross the BBB. Metabolite-laden CSF in SAS on the brain’s superior and lateral surfaces drain to the superior sagittal sinus, where arachnoid granulations facilitate CSF egress. These specialized structures contain unidirectional valves, preventing plasma and blood cells from entering the CNS (Weed, 1914, Welch and Friedman, 1960). However, arachnoid granulations do not form at the brain base, so CSF egress in those areas requires other mechanisms (de Leon et al., 2017, Nedergaard and Goldman, 2020). Recently, glymphatic structures within the meninges have emerged as a potential route for CSF outflow from the basal forebrain of mice (De Mesquita et al., 2018). Another important site for CSF egress from the brain base is the cribriform plate (CP).

The CP, or *lamina cribrosa*, is a porous pocket of ethmoid bone set between the eyes that physically separated olfactory bulbs of the CNS (above) from the nasal cavity (below). Superiorly, the CP’s lateral walls rise to form the olfactory fossae (OF), a paired depression that accommodates olfactory bulbs (OB). Olfactory fossae are separated by a median ridge that is continuous with the nose’s perpendicular bone (below) and crista galli (above). The crista galli is a bulbous structure that rises above the CP to anchors the cerebral falx—a dense connective tissue that separates the frontal lobes up to the superior sagittal sinus. Injuries to the nose can deflect the perpendicular plate and crack the CP, resulting in CSF leak syndrome (rhinorrhea), which lowers intracranial pressure causing headaches and risking encephalitis.

Smells detected by odor receptors in the nasal mucosa are relayed to the brain through cranial nerve 1 (CN1) fibers that bundle together and project upward through CP apertures to glomeruli on the olfactory bulb. Olfactory signals are relayed to the temporal lobe past the basal forebrain, along the olfactory tract and lateral olfactory stria. This contiguous alignment provides an efficient route for CSF drainage from the basal forebrain and medial temporal to the olfactory fossa. Notably, the medial temporal lobe and basal forebrain are the first brain regions affected by AD pathology, including the accumulation of insoluble metabolites, such as fibrillar Aβ, in the interstitial compartment. Advanced age is the most significant risk factor in late-onset AD, and both age and Alzheimer’s disease are associated with dysosmia (Doty et al., 1984; Ottaviano et al., 2016). We previously hypothesized that CP aging reduces CSF flow and metabolite clearance in the medial temporal lobe and basal forebrain, making them susceptible to the accumulation of toxic metabolites (Ethell, 2014).

To explore links between AD and the CP, we used high-resolution imaging and deep learning to characterize CP morphology in AD and control subjects, tested our hypothesis with a new ferret model, and evaluated the CPs of 560 subjects in a clinical study.

## Results

In this project, we undertook three studies to investigate CSF egress through the CP. First, high-resolution micro-CT and deep learning were used to assess ethmoid bone samples isolated from post-mortem subjects, many with a confirmed AD diagnosis. Second, we developed a new ferret model to test the behavioral and pathological effects of CP aperture occlusion. Third, we evaluated CP aging and cognition in a large cohort of volunteers between 20 and 95-years-old (y).

### CSF egress and CP bone

CPs and surrounding bone were isolated from 65 post-mortem subjects (26-94y) and scanned with micro-CT (65,000 gray levels, 16 μm resolution; Fig. 1A). We also scanned the CPs of 560 volunteers with cone-beam CT (3,000 gray levels, 90 μm resolution; Fig 1B). Micro-CT scanning provided exceptional detain of tubular CP apertures and bony conduits (Fig. 1C). While many textbooks describe CP apertures as perforations, we found that was only true on the flat surfaces of the CP floor; the largest apertures occurred along the midline and lateral margins where the bone is much thicker. In those places, apertures formed tubular structures that angled toward the back of the CP as they descended, and those near the front angled anteriorly. Of 624 CPs evaluated in our study, we observed variability in length, width, depth, and pitch that could have influenced aperture formation, size, and distribution (Fig. 1D, E). For example, the subject in C was a 7^th^ decade female (dementia) with a midline asymmetry due to a nose injury that damaged the CP and did not realign before healing. The right side was slightly higher than the left and contained a thickened bony region (arrow) without apertures. In all CPs, the largest apertures ran into the midline ridge, including a distinctive pair at the bottom of the olfactory fossa’s back wall, which projected horizontally into the ethmoid bone, hereafter referred to as ‘jet intakes.’ To automate bone and aperture predictions, we used a fully connected convolutional neural network based on U-Net (Suppl Fig. 1; Ronneberger et al., 2015) trained with manual-segmented micro-CT and cone-beam CT samples. The total cross-sectional area (XAS) of apertures from 330 subjects plotted by age revealed a trendline that declined dramatically around the 7^th^ decade (Fig. 1F). The variability in CP sizes and shapes required an alignment algorithm to normalize CPs and apertures. We then generated aperture probability heatmaps for subjects 55y and under (≤55y; Fig 1G, Suppl Fig. 2) and 65y and over (≥65y; Fig. 1H), then analyzed apertures within the central zone. There were significant differences in the number of apertures between the ≤55y and ≥65y and subjects with a confirmed diagnosis (Fig. 1I) of AD (F(2,265)=32.491, *p*=2.4×10^−13^). Post-hoc analysis revealed no difference in the number of apertures between AD and ≥65, but significant differences between ≤55y and AD (*p*=0.001) or ≥65y (*p*=6.66×10^−14^). These findings establish that age-dependent aperture occlusion is more extensive in AD subjects.

**Fig. 1.**
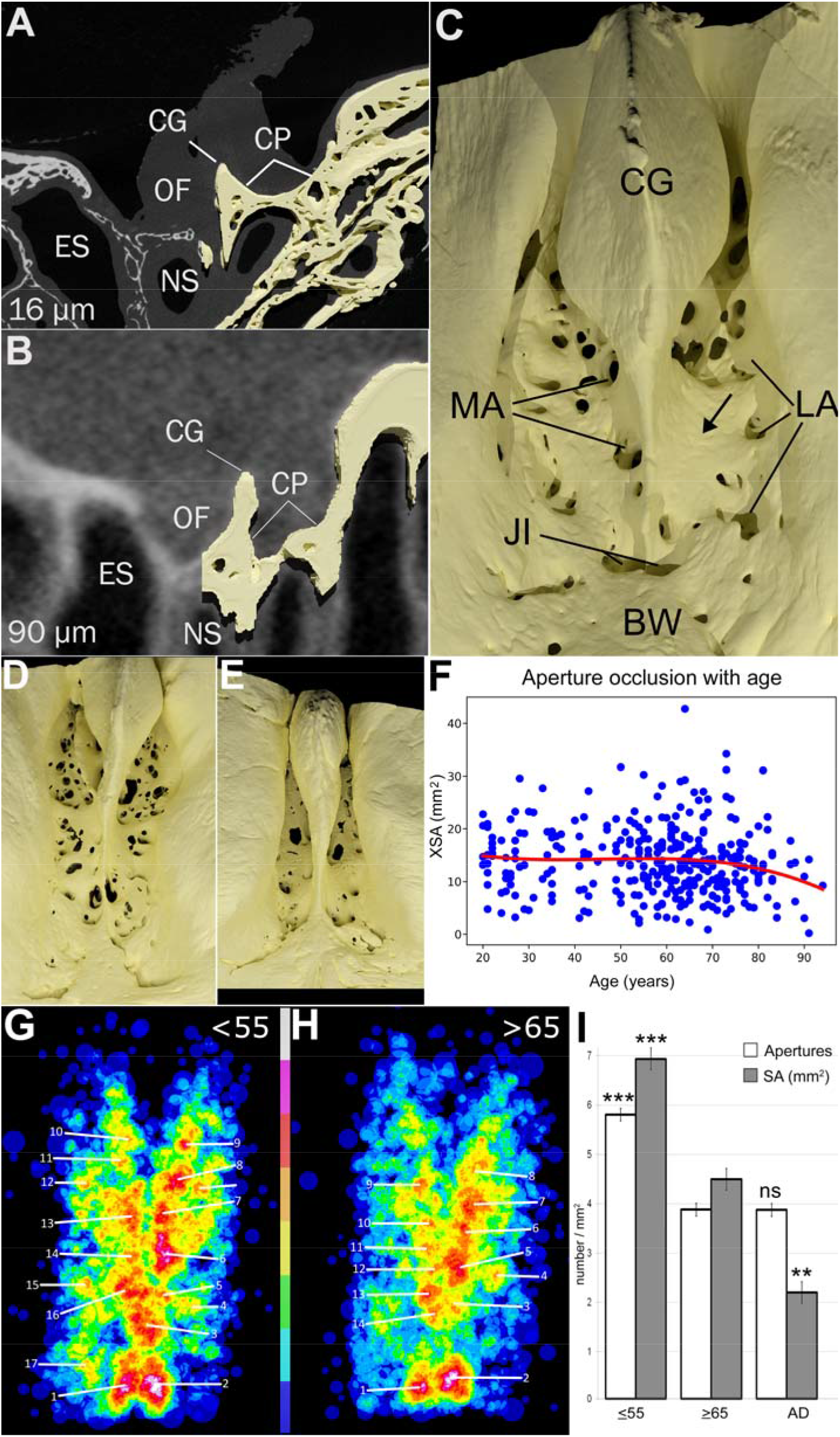
High-resolution anatomy of the human cribriform plate. (**A)** MicroCT scan of an isolated (post-mortem) CP imaged over 2 h at 16 μm resolution with 65,000 gray values, without contrast. The right side of the image is overlaid with bone segmentation from 480 μm of slices toward the viewer (CG, crista galli; CP, cribriform plate; ES, ethmoid sinus; NS, nasal sinus; OF, olfactory fossa). (**B)** Cone-beam CT scan of CP in a live subject imaged over 20 s at 90 μm resolution with 3,000 gray levels, without contrast. The right side of the image is overlaid with bone segmentation from 540 μm of slices toward the viewer (labeled as in A). (**C)** 3D rendering of segmentation of a CP imaged with micro-CT, viewed from above with the front at the top (BW, back wall; CG, crista galli; JI, jet intake apertures; LA, lateral apertures; MA, medial apertures). This CP showed midline asymmetry with the right side sitting higher than the left, with more extensive ossification. The arrow indicates a thickened region of the CP on the raised side that was devoid of apertures. The subject had a confirmed post-mortem diagnosis of AD. Basic CP shapes varied widely from the sample in B, including (**D)** wide bowl-shaped CPs and (**E)** short, narrow, and deep CPs. (**F)** Total cross-sectional area (XSA) of CP apertures in 330 subjects plotted by age (20y-94y). The trendline from all 330 subjects (red) indicates relative stability until the 7^th^ decade after which it declines. (**G)** A Heat map of apertures in CPs of live subjects 55y and younger (≤55), and (**H)** an aperture heat map from subjects 65 and older (≥65). Color scale indicates highest (gray) to lowest (dark blue) probabilities, specific numbers provided Suppl Fig. 2. (**I)** Aperture number and total cross-sectional areas for central apertures in groups ≤55, ≥65, and AD subjects. Asterisks indicate statistical differences to ≥65. Error bars represent standard error of the mean.

Midline CP apertures form angled tubes that connect with conduits in thickened medial bone (Fig. 2A). These conduits run from the crista galli’s cistern to a manifold within ethmoid bone at the olfactory fossa’s back wall, creating a protected watershed between CSF intake from the fossa and the nasal mucosa below (Fig. 2B & inset). This watershed buffers CSF as its flows down a pressure gradient from intracranial pressure (10 mm.Hg) in the olfactory fossa to interstitial pressure in the nasal mucosa (0-2 mm.Hg; Berg et al., 1985) over approximately ∼5 mm. Cone-beams scans consistently showed a lower overall bone density of the ethmoid bone in older subjects, particularly in the lateral walls of the CP. We examined watershed structure in post-mortem subjects and found deterioration with age in many subjects over 65y. However, some AD subjects had relatively intact watersheds, suggesting that watershed capacity is not a rate-limiting step in CSF egress. In contrast, the decrease in total aperture cross-sectional area in AD subjects was significant and represented a more likely choke-point for CSF egress across the CP.

**Fig. 2.**
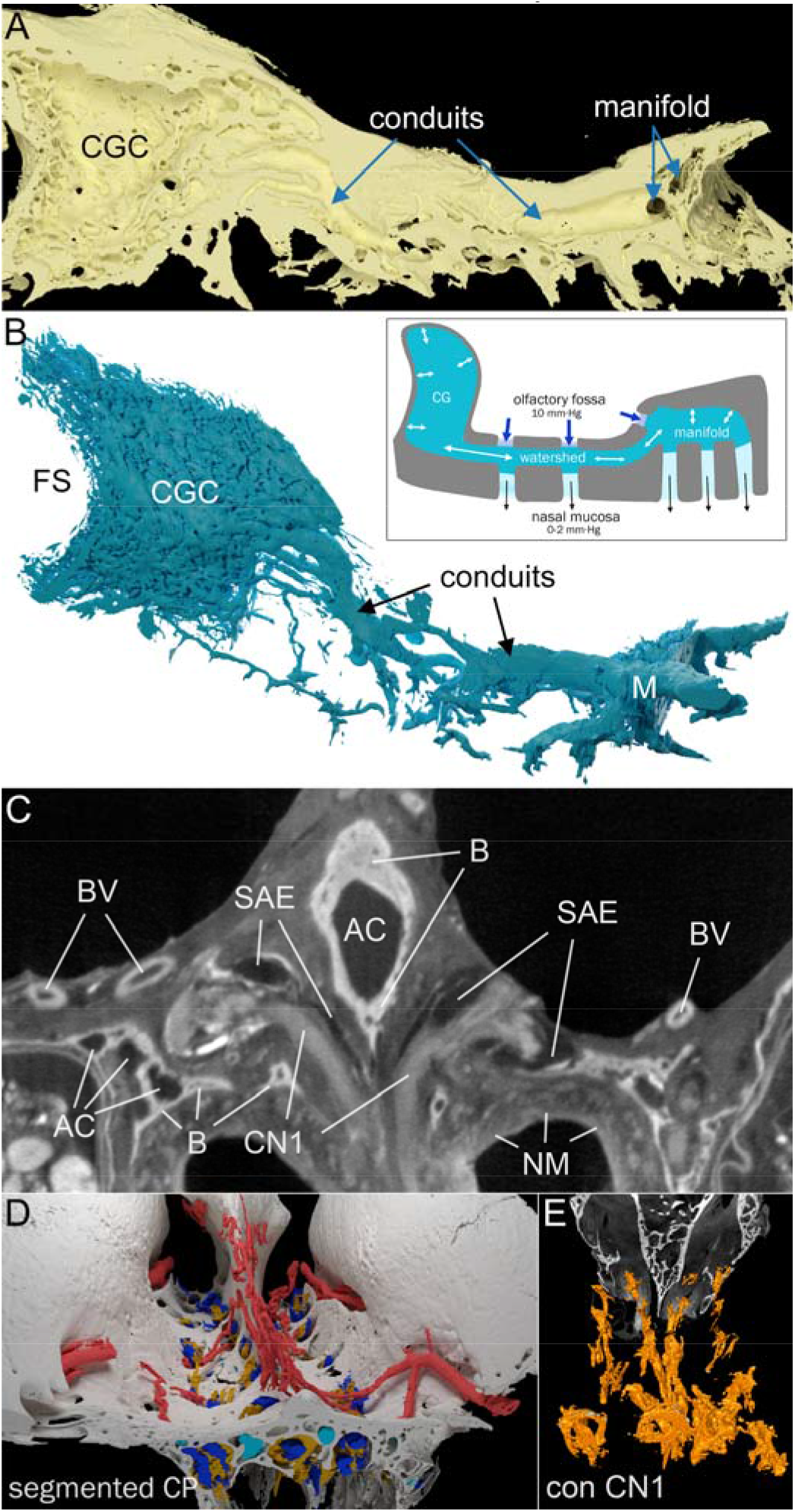
Soft-tissue resolution of the cribriform plate with micro-CT imaging. **(A)** Mid-sagittal cut through a segmented CP from a 4^th^ decade control subject shows the CP’s conduit system running from the anterior crista galli cistern (CGC) on the left to a manifold in the olfactory fossa’s back wall on the right. (**B)** Negative image of the sample in (A) shows the CP watershed from CGC to manifold (M). Anterior to the CGC, a frontal sinus pneumatization (FS) pushes into the CG, but bone separates the CGC from the air in the FS. Image azimuth is shifted upward to the left to show 3D features. (**C)** A coronal micro-CT slice of human CP (4^th^ decade male) with contrast enhancement. Soft tissues avidities for elemental Iodine allow resolution of cranial nerve 1 bundles (CN1), blood vessels (BV), Arachnoid conduit (AC), bone (B), nasal mucosa (NM), and subarachnoid evaginations (SAE). (**D)** Segmentation of a contrast-enhanced micro-CT shows soft tissues in a 3D context. Bone, white; Red, blood vessels; orange, CN1 bundles; blue, subarachnoid evaginations; yellow, bony conduits. (**E)** CN1 nerves from a control subject shown from a forward-facing and elevated angle, with an anterior coronal slice of the micro-CT scan included for orientation.

### CSF egress and CP soft tissues

To resolve soft tissue structures in 3D, we used Iodine-enhanced and micro-CT to image CP bone fragments at 16 μm resolution with 65,000 possible gray levels (Fig. 2C). A spectrum of tissue affinities for elemental Iodine allowed the segmentation of bone, blood vessels, nerves, subarachnoid evaginations, and arachnoid within apertures and bony conduits (Fig. 2D). Significant CN1 losses were apparent in AD subjects compared to age-matched controls (Fig. 2E), consistent with previous reports (Attems et al., 2005). The largest CN1 branches emerged from the jet intakes in the back wall. CN1 deficits in AD subjects were more advanced in anterior apertures, as expected, with OB atrophy retracting them toward the back wall.

With contrast enhancement, liquid-filled areas appeared dark due to the lack of signal, including the crista galli’s cistern (CGC) and subarachnoid spaces (SAS) under the olfactory bulb. We discovered subarachnoid evaginations (SAE) that extended into most apertures, wrapping around CN1 bundles (Fig. 3A, B). Young and middle-aged subjects had SAE in all medial and lateral apertures, with the largest SAE in jet intakes (Fig. 3C). Seniors (≥65) had fewer and smaller SAEs, and AD subjects had the least (Fig. 3D). SAE typically extended halfway down the largest apertures, with branching as they winnowed down.

**Fig. 3.**
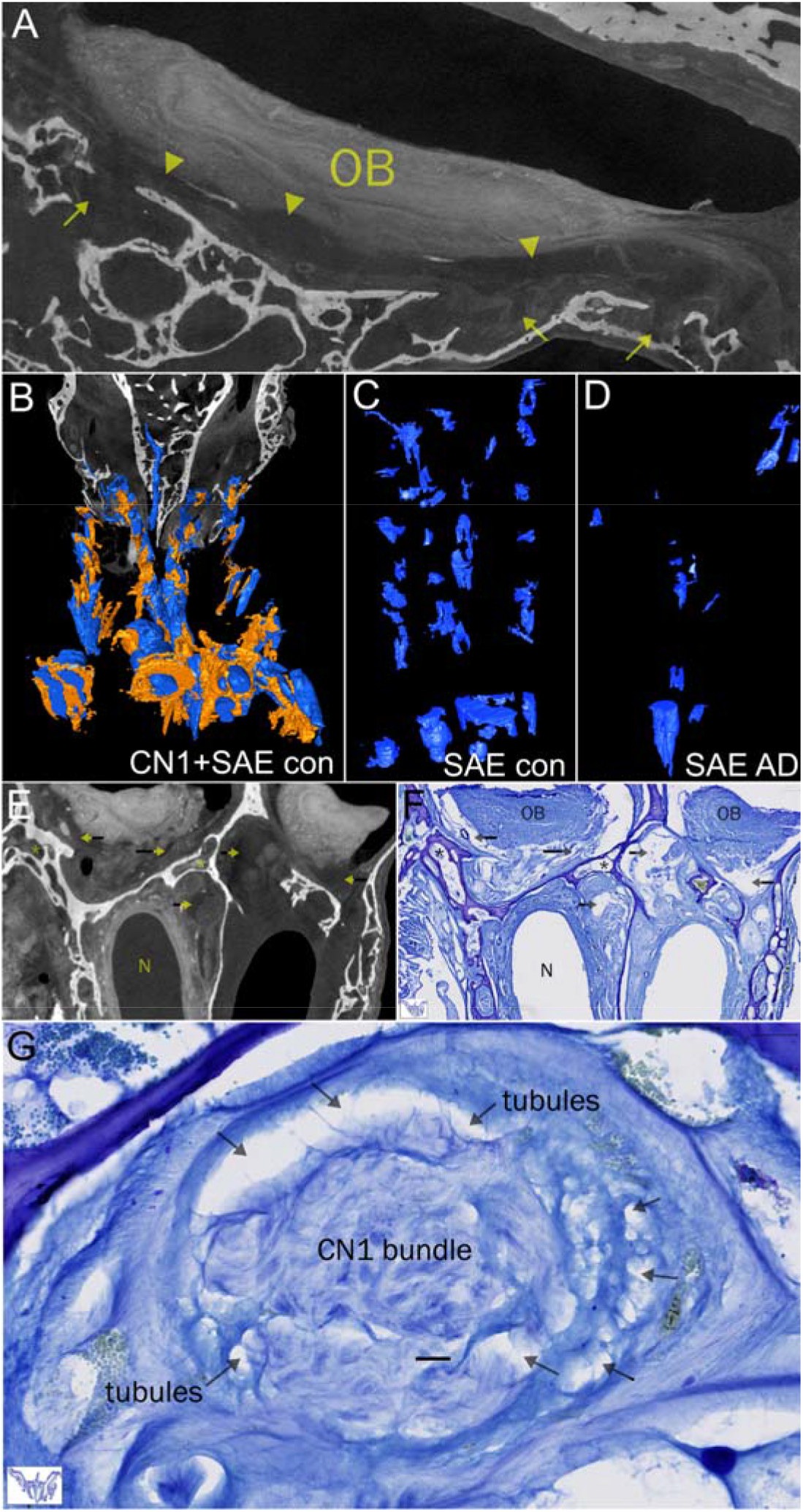
CSF egress through subarachnoid evaginations and aperture tubules. **(A)** Sagittal section through the OB in a human CP that was contrast-enhanced and imaged with micro-CT. Layers and glomeruli in the OB can be seen, as well as CN1 nerves on the right. The dark area below the OB is subarachnoid space (arrowheads) that is continuous with subarachnoid evaginations (SAE) that project down into the apertures (arrows). **(B)** A forward-facing oblique angle of segmented CP with CN1 nerves (orange) and SAE (blue) shown. SAE are intimately connected to CN1 fibers. **(C)** Isolated SAE from a control subject shown from above. SAE from an AD subject from above are greatly reduced compared with (C). **(E)** Coronal slice from a contrast-enhanced micro-CT scan from a control subject, aligned with **(F)** a Nissl-stained 30 μm histological slice in the same position of the sample in (E). Arrows indicate SAS/SAE; OB, olfactory bulb; N, nasal sinus. Scale bar = 100 μm. **(G)** High-resolution image of Nissl staining in an aperture shows CN1 fibers bundled in the middle, surrounded by cuts through tubules (arrows) that surround and intersperse the nerve fibers.

To follow the CSF flow channels further, we sectioned CP samples and stained slices with cresyl violet (Nissl). A comparison of the same section imaged in 3D with contrast-enhanced micro-CT and as a Nissl-stained slice (Fig. 3E, F) revealed SAE structures as they descend into apertures (Fig. 3G). Arachnoid cells under the SAS are continuous with cells that form SAE and tubules. Smaller tubules were similar to the spider-like cores of arachnoid granulations in the superior sagittal sinus (Kida et al., 1988), at the top of the cerebral falx, and extended down to the nasal mucosa. Importantly, tubules also branched into the conduit system, which contained arachnoid cells arranged in a porous configuration around a central open channel. This channel provides rapid dissipation of pressure differentials along the watershed’s length from crista galli to manifold. As with arachnoid granulations, the SAE, tubules, and watershed favor unidirectional CSF flow, from olfactory fossa to nasal mucosa. As CSF enters the top of a CP aperture, its pressure drops from intracranial pressure (∼10 mm.Hg) to a theoretical intermediate pressure (TIP) stabilized by a larger CSF volume in the watershed, including the CGC and manifold. CSF continuing down an aperture from the watershed loses pressure again until it reaches the nasal mucosa. This arrangement counters backflow from increases in the interstitial pressure of nasal pressure due to position or sneezing, creating a one-way system that prevents pathogen entry to the brain.

Per Bernoulli’s principle, the total aperture cross-sectional area of CP apertures restricts flow capacity and hence CSF egress. Age-dependent declines in flow capacity disrupt CSF egress through the CP, resulting in less CSF drainage from upstream brain regions, including the basal forebrain and medial temporal lobe. These findings support our hypothesis that age-dependent declines in CSF egress across the cribriform plate seed AD pathology by allowing the accumulation of interstitial metabolites, including Aβ oligomers (Ethell, 2014).

## Experimental aperture occlusion

Out pathological analysis of post-mortem CPs showed a significant correlation between CSF egress capacity and AD. To provide conclusive evidence of a cause-and-effect relationship, we tested our hypothesis with a new animal model. Unlike rats and mice, Ferrets are curious and intelligent carnivores with the same Aβ1-42 amino acid sequence as humans. As in all quadrupeds, the cribrose structure of ferrets is vertically oriented, with apertures divided into upper and lower regions. Approaching the nasal side, we surgically occluded the upper CP apertures in adult ferrets with dental cement, reducing CSF flow capacity by half. This procedure was done through a small window in the upper snout, which had no overt impact on movement, behavior, or socialization for at least several weeks post-op. Occluded (n=8) and control ferrets (n=4) were housed together and evaluated for 7-months post-op. Spatiotemporal memory (STM) was evaluated in a tube maze that required 2 or 3 correct turns to solve. Each testing day consisted of five maze run trials for each animal, and we used T4 and T5 as indicators of their ability to learn the maze. One month before surgery, ferrets were acclimated to the test with a one-decision point maze (1-DPM). After allowing one month for a full recovery from surgery, ferrets were run through a 2-DPM every two weeks for 2-months, with no significant differences observed between the two groups. As the ferrets were proficient in the 2-DPM at 2-months, we increased the difficultly by adding one more decision point (3-DPM), swapping between all-left turns and all-right turn DPMs on biweekly testing days. Differences between the two groups emerged after two months, with occluded ferrets showing a progressive decline in maze solving ability (Fig. 4A).

**Fig. 4.**
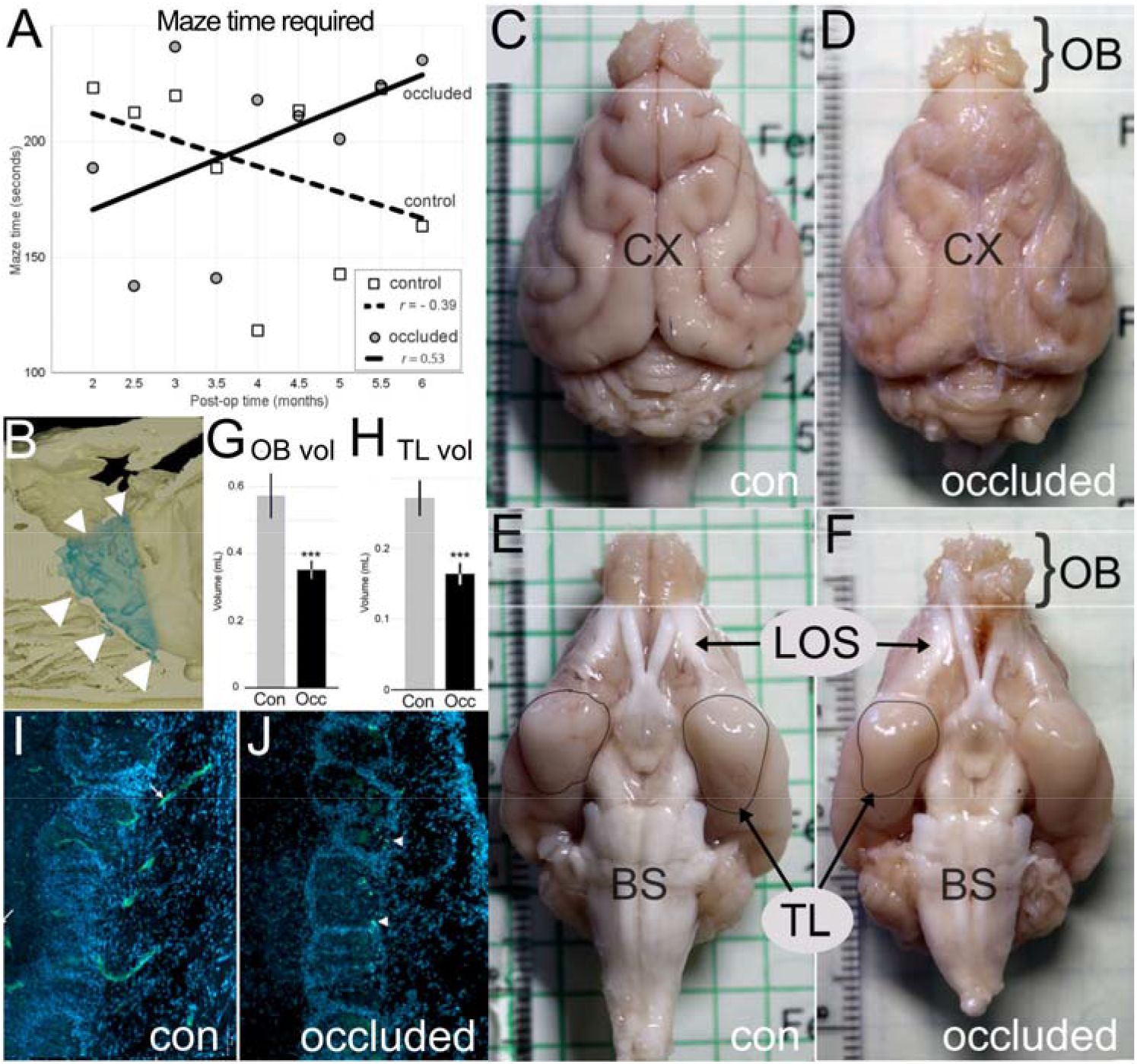
Surgical blockage of aperture induces atrophy and cognitive deficits. **(A)** Biweekly spatiotemporal memory performance of control and CP occluded ferrets in a tube maze with three decision points. Control ferrets (square) became faster in the maze up to 6.5 months post-op, with a negative trendline (dashed line; *r* = -0.39, *p*=0.293); whereas, CP occluded ferrets (gray circle) became progressively worse with at the maze, requiring more time to complete it with a trendline (solid black) in the opposite direction (*r* =0.54, *p*=0.146). **(B)** Mid-sagittal cut through the skull (segmented) of a CP occluded ferret, front of the head is to the left. The location of the OB is shaded blue. Nasal turbinates outside the upper apertures (small arrowheads) were removed and replaced with a dental cement plug (not shown). Lower apertures (large arrowheads) connect to intact turbinates. **(C)** Dorsal views of brains from a control ferret, and **(D)** a CP-occluded ferret, 7-months post-op. Olfactory bulbs (OB) are at the top, indicating the start and end of OB in control ferrets. The occluded ferret brain had extensive atrophy of the OB. **(E)**Ventral views of the same two brains showing the extent of OB atrophy as well as atrophy of the lateral olfactory stria (LOS) and **(F)** temporal lobe (TL). The temporal lobe (aka piriform lobe) was outlined in both samples. Compare smoothing of the occluded ferret’s temporal lobe (left side of the image) to the corresponding region in the control brain. **(G)** Bar chart of OB and **(H)** temporal lobe volumes in all control and occluded ferrets showing significant volume reductions in the ferrets with cribrose aperture blockade. **(I)** Comparison of DAPI+thioflavin S-stained nuclei in control and **(J)** occluded ferret OB show depletion of cells around the glomeruli and thioflavin-positive inclusions after cribrose aperture blockade. Thioflavin (green) staining in the control brain (J) was due to vascular structures.

We also noticed changes in the demeanor of occluded ferrets that suggested reduced anxiety and fear. For instance, testing days involved ferrets running through the same maze five times, with rest periods in-between. By the third or fourth test, control ferrets were irritated with the task and tried escape handlers with squirming and biting. However, by the 3^rd^-4^th^ month, occluded ferrets were no longer aggressive but placid with no struggling (i.e., they went limp like a friendly cat). We observed this loss of aggression in 7/8 occluded ferrets, although one outlying ferret responded to handling like the control animals. Further, when control ferrets became frustrated with the maze in T1-T3, they would sometimes vocalize distress squeaks, which were not made by occluded ferrets, beyond 3-months post-op. Indeed, after they timed out, occluded ferrets were sometimes found lying on their backs calmly playing with the maze walls.

Seven months after surgery, all ferrets were sacrificed and perfused with paraformaldehyde for pathological analysis. Ferret heads were imaged with micro-CT (Fig. 4B), and the brains were removed by careful dissection, taking care not to damage brain tissue. Isolated brains were photographed from dozens of angles (200-400 images) and used to render a digital 3D representation to calculate regional brain volumes (Fig. 4C-F). Occluded ferrets had significantly (*p*=0.0013) smaller OBs (0.351 ± 0.026 mL) than control ferrets (0.573 ± 0.065 mL), representing a 39% loss of OB volume (Fig. 4G). The temporal lobes of occluded ferrets were smaller (0.164 ± 0.024 mL) compared with control ferrets (0.272 ± 0.014 mL; Fig. 4H), and they also had thinner lateral olfactory stria (Fig. 4E, F).

DAPI-stained OB sections showed fewer nuclei around glomeruli in occluded versus control ferrets (Fig. 4I, J) and the appearance of numerous thioflavin S deposits in the glomerular layer (Fig. 4K, L).

## Correlations between cognitive decline and CP occlusion

We undertook a clinical study, Project Cribrose, to investigate relationships between CP morphology, cognition, olfaction, and clinical variables. A large and diverse population of 560 volunteers was enrolled from Riverside, California, USA. After providing informed consent, participants answered a questionnaire about their age, sex, ethnicity, smoking, education, occupation, head and facial injuries, family history of dementia, and more. A cognitive testing platform (LMT) was developed to produced multi-dimensional data suitable for machine learning, including tests for spatiotemporal memory, semantic memory, pattern recognition, processing speed, word recall, and alternating tasks. LMT required 15-20 min to complete using a touchscreen computer that recorded responses and latencies, encrypted the data, and transmitted it to an internal server. Each LMT component showed age-dependent deficits. Minor progressive, and synchronous deficits, occurred in object recall (*R*^2^ =0.876, *p*=0.0044), STM (*R*^2^ =0.864, *p*=0.0003), and pattern recall (*R*^2^ =0.902, *p*=0.0022) until the end of the 6^th^ decade, after which those tasks became more difficult (Fig. 5A). Deficits in word recall were progressive between 20y and 94y (*R*^2^ =0.976, *p*=3.39×10^−5^), but errors in an alternating task (*R*^2^ =0.921, *p*=0.0011) increased after the 7^th^ decade (Fig 5B). Lastly, processing speed did not deteriorate until the 8^th^ decade (*R*^2^ =0.867, *p*=0.0025).

**Fig. 5.**
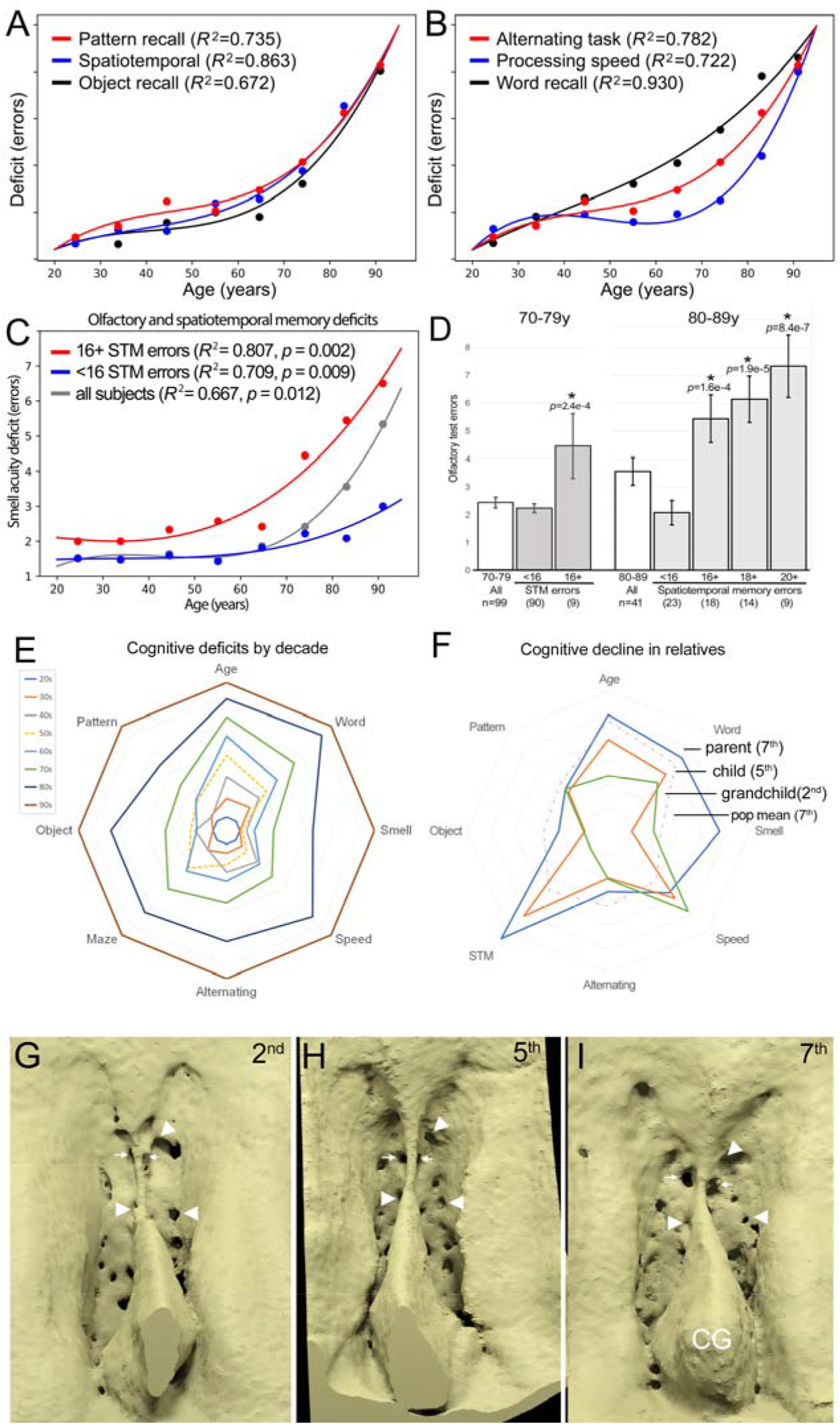
Synchronous decline of cognition, smell, and aperture size. **(A)** LMT results plotted by age, grouped by decade, for pattern recall errors (red), maze memory errors (Spatiotemporal, blue), and object recall errors (black). **(B)** LMT results for Alternating task score (red), processing speed score (blue), word recall errors (black). R^2^ values listed in A and B; *p*-value and *r* provided in the text. **(C)** Olfactory discrimination made by all subjects by age, grouped by decade (dotted line). Subjects with 16 or more errors in the STM test (red line), subjects with less than 16 STM errors (blue line), and all subjects (gray line). Note divergence between groups based on STM errors after the 6^th^ decade. **(D)** Statistical analysis of the STM errors and olfactory errors in the 7^th^ and 8^th^ decades. Subjects with 16+ STM errors made significantly more smell test errors in both decades. Numbers of smell test errors and STM errors (16+, 18+, 20+) made in the 8^th^ decade were highly correlative. **(E)** Radar plot of decade means for LMT, smell, and age, normalized between 25 and 95y. Youngest subjects in the middle with fewest errors on all tests, with successive decades spreading outward as concentric rings. Color code key provided. 5^th^ decade contrasts with a dashed line as it crossed 4^th^ and 6^th^ decades in some measures. **(F)** Radar plot of LMT and smell results for a parent (7^th^ decade, blue), child (5^th^ decade), and grandchild (2^nd^ decade, green). Progressive results culminated in the parent displaying deficits in STM, word recall, object recall, and smell that exceeded population means for 7^th^ decade (dashed gray). **(G-I)** Similarities between the cribriform plates of these three subjects illustrate correlations between cognitive decline and CP aging. The back wall is to the top, and jet intakes are indicated with arrows. There is some variability in apertures, but examples of apertures that occlude are indicated with arrowheads. The 2^nd^ decade grandchild (G) had large well-defined apertures, although some apertures were obscured by bone. Crista gallies in G and H were cut-off because they extended higher than the predicted ROI. The 5^th^ decade child (H) had a deeper olfactory fossa and smaller apertures than her child. Some bone-thinning is indicated by a roughened appearance of this bone prediction. The parent (I) showed aperture occlusion with smaller and fewer apertures as well as bone-thinning. Note the top arrowhead indicates an occluded aperture. This 7^th^ decade individual also showed enlargement of the crista galli (CG).

Pearson’s *r* showed significant correlations between the decline of all LMT components (Table 1) from the lowest correlation between word recall and processing speed (*r*=0.89, *p*=0.003) to the highest between processing speed and smell acuity (*r*=0.99, *p*=7.9×10^−7^). We divided participants into 3 three age groups, ≤55y, 56-64y, and ≥65y, and performed one-way ANOVA, which showed significant between-group differences in all LMT components and olfaction. Paired Post-hoc tests with Bonferroni correction showed significant differences between ≥65y and ≤55y groups for each LMT component (Table 2).

**Table 1.** Pearson correlation coefficient values for LMT and smell test results.

|  |  | Smell | Word | STM | Alt | Speed | Pattern | Object |
| --- | --- | --- | --- | --- | --- | --- | --- | --- |
| Word errors | $r$ | 0.903 | | | | | | |
| | $R^2$ | 0.815 | | | | | | |
|  | T-statistic | 5.15 |  |  |  |  |  |  |
| | $p$-value | 0.0021 | | | | | | |
| STM errors | $r$ | 0.964 | 0.971 | | | | | |
| | $R^2$ | 0.930 | 0.944 | | | | | |
|  | T-statistic | 8.90 | 10.02 |  |  |  |  |  |
| | $p$-value | $1.12 \times 10^{-4}$ | $5.74 \times 10^{-5}$ | | | | | |
| Alt score | $r$ | 0.964 | 0.978 | 0.980 | | | | |
| | $R^2$ | 0.930 | 0.957 | 0.961 | | | | |
|  | T-statistic | 8.93 | 11.53 | 12.19 |  |  |  |  |
| | $p$-value | $1.1 \times 10^{-4}$ | $2.55 \times 10^{-5}$ | $1.86 \times 10^{-5}$ | | | | |
| Speed score | $r$ | 0.993 | 0.890 | 0.957 | 0.954 | | | |
| | $R^2$ | 0.986 | 0.792 | 0.915 | 0.910 | | | |
|  | T-statistic | 20.86 | 4.78 | 8.05 | 7.77 |  |  |  |
| | $p$-value | $7.90 \times 10^{-7}$ | 0.0031 | $1.96 \times 10^{-4}$ | $2.39 \times 10^{-4}$ | | | |
| Pattern errors | $r$ | 0.947 | 0.940 | 0.962 | 0.957 | 0.947 | | |
| | $R^2$ | 0.896 | 0.884 | 0.925 | 0.915 | 0.896 | | |
|  | T-statistic | 7.21 | 6.77 | 8.58 | 8.04 | 7.21 |  |  |
| | $p$-value | $3.61 \times 10^{-4}$ | $5.07 \times 10^{-4}$ | $1.38 \times 10^{-4}$ | $1.97 \times 10^{-4}$ | $3.61 \times 10^{-4}$ | | |
| Object errors | $r$ | 0.976 | 0.950 | 0.990 | 0.979 | 0.975 | 0.941 | |
| | $R^2$ | 0.953 | 0.902 | 0.980 | 0.958 | 0.951 | 0.886 | |
|  | T-statistic | 10.99 | 7.42 | 16.98 | 11.72 | 10.84 | 6.84 |  |
| | $p$-value | $3.38 \times 10^{-5}$ | $3.08 \times 10^{-4}$ | $2.66 \times 10^{-6}$ | $2.32 \times 10^{-5}$ | $3.66 \times 10^{-5}$ | $4.80 \times 10^{-4}$ | |
| Aperture XSA | $r$ | -0.941 | -0.782 | -0.854 | -0.866 | 0.962 | -0.901 | -0.875 |
| | $R^2$ | 0.885 | 0.612 | 0.729 | 0.75 | 0.925 | 0.812 | 0.766 |
|  | T-statistic | 4.65 | 11.56 | 15.92 | 16.96 | 36.32 | 20.85 | 17.84 |
| | $p$-value | 0.0035 | $2.52 \times 10^{-5}$ | $3.90 \times 10^{-6}$ | $2.68 \times 10^{-6}$ | $2.90 \times 10^{-8}$ | $7.92 \times 10^{-7}$ | $2.00 \times 10^{-6}$ |

n=8 df=6 2-tail

**Table 2.**
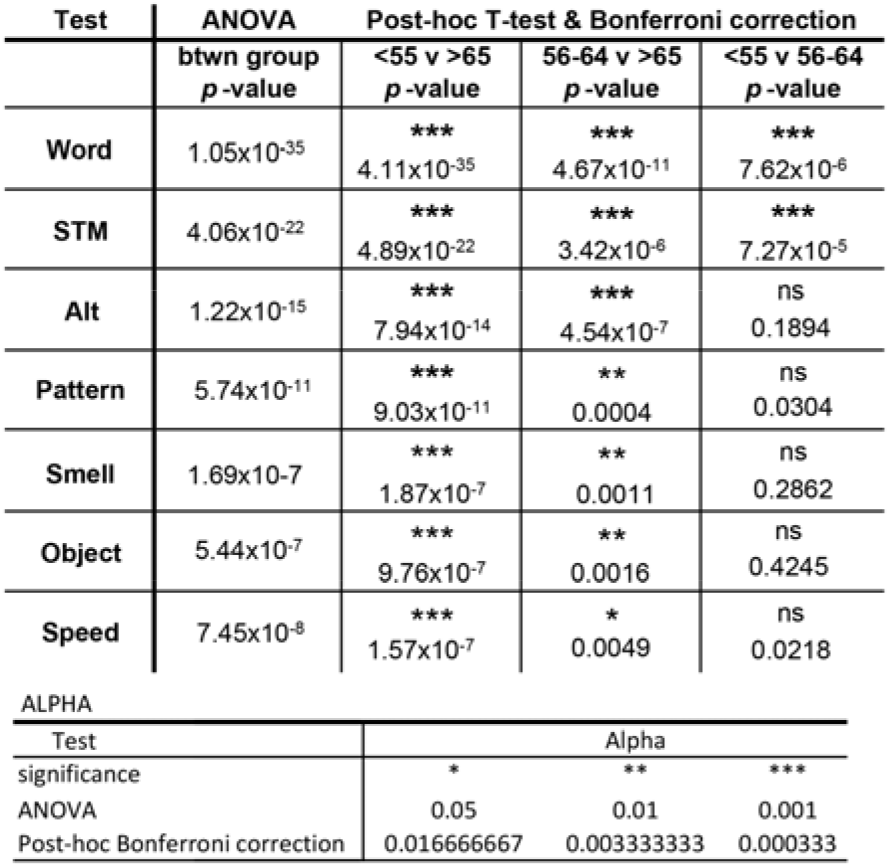
One-way ANOVA comparing LMT and smell test results in age groups: ≤55y, 56-64y, and ≥65y.

Smell acuity was evaluated with a 10-query test that involved removing a sticker to uncover an absorbed drop of essential oil on a card. Subjects sniffed the spot and selected from four possible smells printed on the card. Testing revealed a progressive loss of olfactory discrimination between 20y and 90y (*R*^2^=0.680, *p*=0.0117; Fig. 5C), consistent with previous reports (Doty et al., 1984). Pearson correlation coefficients between smell and LMT tests (Table 2) were all significant, ranging from word recall (*r*=0.90, *p*=0.002) to processing speed (*r*=0.99, *p*=7.90×10^−7^). Deficits in spatiotemporal memory and olfaction are early features of AD and progressed synchronously in our study population, so we used one-way ANOVA to compare those two measures (Fig. 5D). There were no significant differences in smell identification between young and middle-aged subjects (under 60y) with poor STM compared to good STM. However, subjects in their 7^th^ and 8^th^ decades that performed poorly on the STM test were significantly worse at smell identification (*p*= 2.4×10^−4^ and *p*=1.3×10^−4^, respectively). For example, post-hoc (paired T-tests) analyses with the Bonferroni correction determined that subjects in their 8^th^ decade that made at least 16 STM errors made significantly (*p*=1.6×10^−4^) more olfactory test errors (5.4±0.86, 6.14±0.95, 7.33±1.12) than subjects with <16 STM errors 2.09±0.35. Radar plots of LMT and smell deficits revealed progressive losses from the 2^nd^ through 9^th^ decades (Fig. 5E). We used quadratic regression to predict theoretical scores for each subject, based on age, and compared with their testing scores.

To illustrate inherited effects on CP morphology and age-dependent ossification, we present three women from one family with similar faces and cribriform plates, including a parent (7^th^ decade), child (5^th^ decade), and grandchild (2^nd^ decade; Fig. 5F-I). A radar plot of LMT and smell testing shows a progression of deficits in STM, word recall, and object recall, with deterioration of smell acuity that accelerates in the 7^th^ decade (Fig. 5F). All three share facial features around the bridge of the nose and eyes, as well as structural features in their CPs, including aperture patterns that illustrate occlusion (Fig. 5G-I).

## Discussion

We characterized CSF egress through human CP and found similarities to arachnoid granulations with the addition of a watershed that provides constant outward flow and buffers pressure fluctuations on either side of the CP. The choke point for CSF egress is aperture intake at the olfactory fossa, calculated as the total aperture cross-sectional area, which declines with age, becoming extreme in AD. Using a new animal model, we demonstrated that CP aperture occlusion induces cognitive impairment and atrophy of brain areas impacted by early AD. These findings provide pivotal evidence that CSF-mediated clearance is critical for brain homeostasis and suggest that tracking CP apertures may be a valuable indicator of AD risk and cognitive decline.

Many investigators have reported dysosmia in AD patients (Rezek, 1987), and here we provide a mechanism that explains that link. Age-dependent aperture occlusion also impacts the olfactory system’s regenerative capacity. Odor receptors in the nasal mucosa regenerate every 2-3 weeks, and newly formed receptors send axonal growth cones to the OB by fasciculation with older CN1 processes in CP apertures (Schwob et al., 2017). Unmyelinated CN1 fibers and arachnoid-structured tubules share finite space within the confines of a tubular CP aperture, and it is unclear which is most sensitive to aperture closure. Dysosmia and declining STM were significantly correlated in subjects over 70y (Fig. 5A-D) but not before, which may reflect other causes of dysosmia, such as hyperthyroidism (McConnel et al., 1975), before the 7^th^ decade. Ferrets showed extensive OB atrophy, with minimal sparing of glomeruli, seven months after partial occlusion of their cribrose structure (Fig. 4B-F). While the lack of target-field innervation could account for most of the loss, the degree of OB degeneration was exceptional.

These findings add to growing evidence that CSF flow and egress play central roles in brain homeostasis (Nedergaard and Goldman, 2020). Metabolite-laden CSF drains into the olfactory fossa and flows continuously from SAS to SAE as they descend into tubular apertures (Fig. 2-3). SAE subdivide into smaller tubules that carry CSF to the nasal cavity and exchange CSF with the CP’s watershed. The CP watershed is an interface between high intracranial pressure in the olfactory fossa (above) and low interstitial pressure in the nasal mucosa (below), enabling it to buffer pressure fluctuations and maintain unidirectional CSF flow (Fig. 1H, I). Tiny tubules at the bottom of apertures combined with the constant outward flux of CSF present a significant barrier to entry for motile pathogens in the nasal mucosa that protects the brain from infection. Further, antimicrobial peptides in CSF, including Aβ (Soscia et al., 2010), neutralize and trap adherent pathogens and flush them back to sinus tissue. Our discovery of the CP’s watershed adds a new function to the crista galli—a peculiar bony structure that until now has been considered a passive anchor for the cerebral falx. The CSF volume in the crista galli’s cistern provides the watershed with the capacity to buffer a 10 mm drop in pressure over ∼0.5-2 mm. Our ferret study established that reducing CSF egress at the CP could induce cognitive changes similar to mild cognitive impairment. Ferrets with only half of their apertures occluded showed progressive memory deficits and loss of fear, consistent with the hippocampus and amygdala dysfunction, respectively. Our clinical study documented significant correlations between cognitive decline and CP occlusion in subjects ≤55y and ≥65y.

The age at which aperture occlusion reaches a critical level depends on environmental and genetic factors. We inherit facial features from our parents and grandparents, including bony elements such as the CP and ethmoid bone (Fig. 5F, G). While studying >600 human CPs, we encountered significant variability in length, width, depth, and pitch that could impact aperture formation, size, distribution, and probably occlusion rate. Monogenic mutations in PSEN1, or PSEN2, are linked with most early-onset AD cases, but polygenic influences on CP morphology could also predispose some families to aperture occlusion and increased risk for AD or frontotemporal dementia. Down syndrome (trisomy 21, DS) patients typically have brachycephaly (“pug-nose”) that impacts CP morphology, along with life-long cognitive impairment and a predisposition to AD-like pathology beginning in the 3^rd^ decade (Lott, 2012). AD pathology in DS patients is attributable to gene-dosage effects of APP on chromosome 21. However, post-pubescent DS patients develop dysosmia/anosmia (Ceccini et al., 2016), and adults have dilated temporal horns of the lateral ventricles (Pearlson et al., 1998). CP aperture blockage could contribute to overall cognitive impairments in DS at all ages and warrants investigation. Environmental risks to CP integrity include head trauma and injuries that deflect the nose as the septal cartilage is continuous with the perpendicular bone and the CP’s medial ridge. We found that extreme asymmetries often included more extensive aperture occlusion on one side (Fig. 5G), suggesting that early-life injuries may alter CP occlusion over decades. This reasoning could explain elevated AD risk associated with traumatic head injuries (Hof et al., 1992) and cognitive disorders in boxers and professional football players (Alosco et al., 2017). Furthermore, transient losses of olfaction in many COVID-19 patients and cognitive deficits in Long-COVID patients (Sudre et al., 2021) may be related to localized inflammation that impairs CSF egress through the CP.

Recent advancements in deep learning provide new opportunities to glean mechanistic insights from anatomy and population studies. For example, 3D-UNet allowed us to train models with extremely high-resolution samples from post-mortem micro-CT samples and then use those models to predict bone patterns from clinical CT scans with lower resolution. The cone-beam CT scanner we used produces 3,000 gray levels, but the human eye distinguishes less than 20 gradations, making hand-segmentation difficult and time-consuming. However, ML algorithms treat voxels as normalized floating-point calculations, so combinations of voxels that denote edges and boundaries in a 65,000 gray level scan can be normalized to predict bony structures in a cone-beam CT with lower resolution. Our study pioneers a new approach to studying human tissues and disease by incorporating cutting-edge deep learning, an advanced method for ultra-resolution contrast-enhanced micro-CT, a novel animal model, and a large clinical study designed to produce highly curated data for machine learning. We used MIMD-DL to combine imaging, categorical, and numerical data into prognostications that may presage cognitive impairments years in advance. A hybrid approach could also incorporate massive data troves collected by mobile devices and digital monitors to provide a holistic understanding of when disease processes begin, aiding the development of earlier interventions. For example, multi-input mixed data (MIMD) models can integrate categorical, quantitative, and imaging volumes to drive and test hypothesis-based research. This technology represents an epochal shift in biology and medicine that will allow us to develop new ideas and re-evaluate what is ‘known’ about human anatomy using activity, behavior, gross anatomy, tissue, cellular, genetic, and epigenetic markers.

Our characterization of CSF egress mechanisms in the CP has implications for diagnosing, treating, and preventing neurological conditions. It provides conclusive evidence that CP aging plays a critical role in Alzheimer’s disease etiology.

## Data Availability

Relevant data and information are provided in the supplementary information.

## Acknowledgments

We thank Tea Jashashvili for assistance with micro-CT imaging, Amy Chew for anatomical dissections, David Wolf and Diane McClure for animal model selection, Zoe Figueroa and Mary Hamer for technical assistance. Randall Woltjer provided CP samples from post-mortem subjects and neuropathological summaries. Funding was provided by Leucadia Therapeutics Inc. and The Methuselah Foundation. We thank donors to The Willed Body Program at WesternU and their families for their generous contribution to scientific research.

## Funding

Funding provided by Methuselah Foundation and Leucadia Therapeutics Inc.

## Author contributions

Conceptualization: DWE, DM, JS

Methodology: DWE, RZ, DM, TC, SK

Investigation: DWE, RZ, DM, TC, SK

Visualization: DWE, RZ

Funding acquisition: DWE

Project administration: DWE, AO, DM

Supervision: DWE

Writing – original draft: DWE

Writing – review & editing: AO

## Competing interests

DWE, RZ, DM, and JS hold equity in Leucadia Therapeutics Inc. DWE holds multiple patents. AO, SK, and TC declare they have no competing interests.

## Data and materials availability

Code will be provided on GitHub. Imaging files will be made available for non-commercial use under a signed material transfer agreement with Leucadia Therapeutics Inc.

## Supplementary Materials

Materials and Methods

Figs. S1 to S2

Tables S1

## Notes

### Competing Interest Statement

DE, RZ, DM, and JS hold equity in Leucadia Therapeutics. AO, SK, and TC declare no competing interests

### Clinical Trial

Clinical data collected was observational and non-interventional.

### Funding Statement

Funding was provided Methuselah Foundation and Leucadia Therapeutics Inc.

### Author Declarations

An observational clinical study was done under an IRB protocol approved by the Arrowhead Regional Medical Center IRB Committee. An IRB to harvest and study human cribriform plate tissue samples was approved by the Western University of Health Sciences IRB Committee. An animal study was done under an approved IACUC protocol from Loma Linda University's IACUC Committee.

