## Supplementary figures and images for "Impairment of CSF Egress through the Cribriform Plate plays an Apical role in Alzheimer’s disease Etiology"

### Supplemental Figure 1

**Supplemental Figure 1. Modified U-Net setup**

Zaragoza et al., 2021.

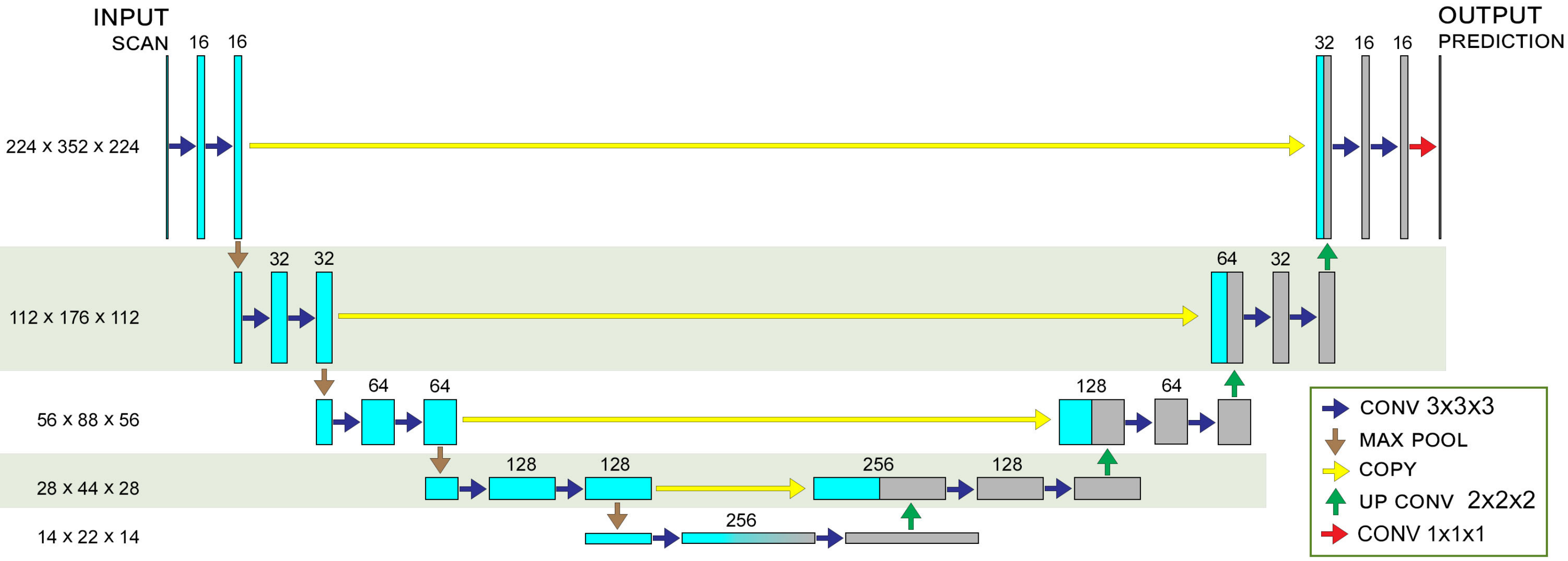

### Supplemental Figure 2

Supplemental Figure 4. Three decision point tunnel maze (3-DPM)

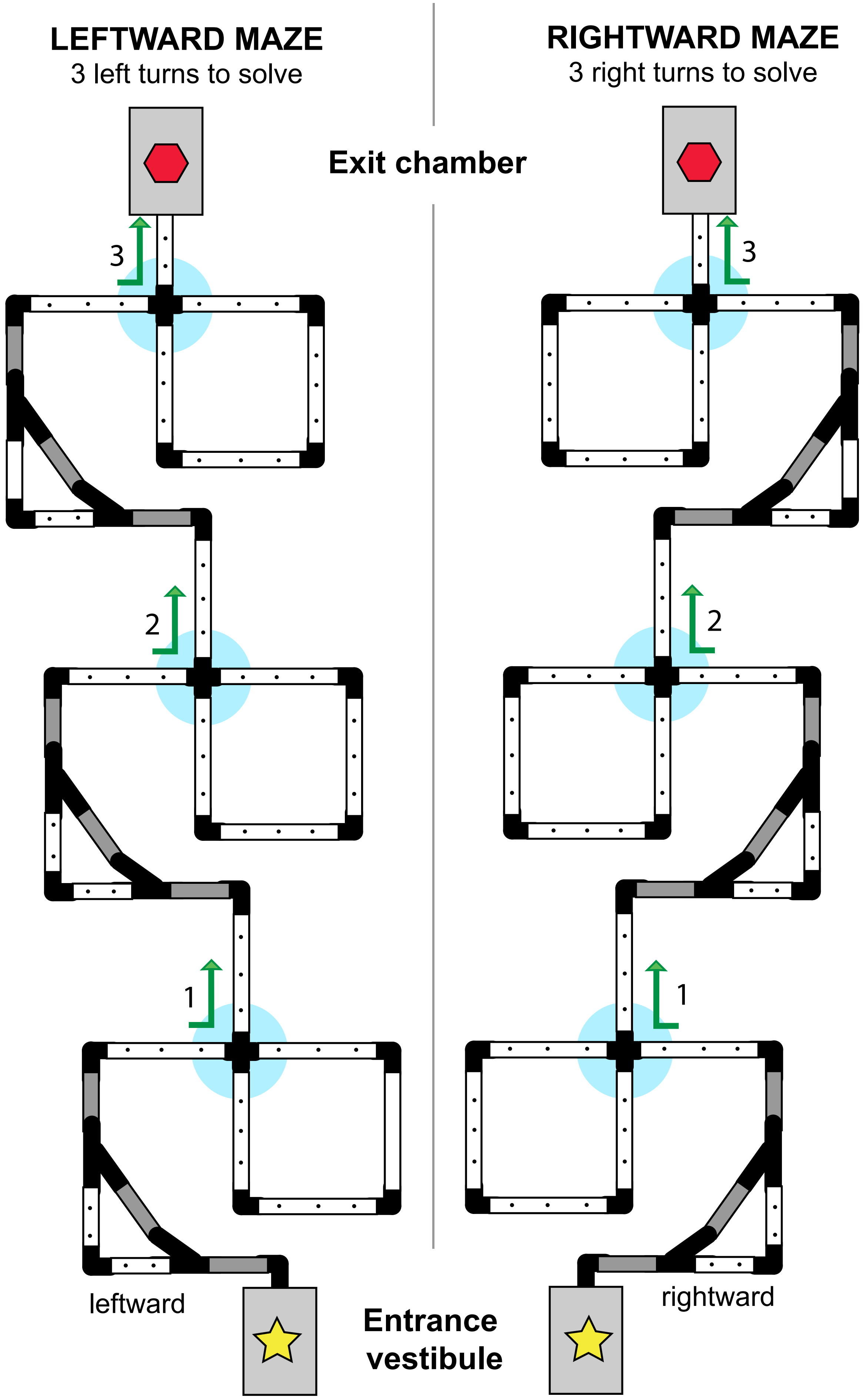


Zaragoza et al., 2021.
