## Supplemental Table 1 for "Impairment of CSF Egress through the Cribriform Plate plays an Apical role in Alzheimer’s disease Etiology"

Supplemental Fig. 2. Aperture heat map probabilities for subjects $\leq$55 and $\geq$65

**$\leq$55 y**

| Heatmap Location | Probability | Percentage |
|---|---|---|
| 1 | 0.734 | 73.4 |
| 2 | 0.726 | 72.6 |
| 3 | 0.411 | 41.1 |
| 4 | 0.129 | 12.9 |
| 5 | 0.21 | 21 |
| 6 | 0.621 | 62.1 |
| 7 | 0.323 | 32.3 |
| 8 | 0.339 | 33.9 |
| 9 | 0.266 | 26.6 |
| 10 | 0.145 | 14.5 |
| 11 | 0.137 | 13.7 |
| 12 | 0.226 | 22.6 |
| 13 | 0.556 | 55.6 |
| 14 | 0.105 | 10.5 |
| 15 | 0.161 | 16.1 |
| 16 | 0.565 | 56.5 |
| 17 | 0.153 | 15.3 |

**>65**

| Heatmap Location | Probability | Percentage |
|---|---|---|
| 1 | 0.463 | 46.3 |
| 2 | 0.559 | 55.9 |
| 3 | 0.154 | 15.4 |
| 4 | 0.132 | 13.2 |
| 5 | 0.471 | 47.1 |
| 6 | 0.265 | 26.5 |
| 7 | 0.456 | 45.6 |
| 8 | 0.162 | 16.2 |
| 9 | 0.213 | 21.3 |
| 10 | 0.191 | 19.1 |
| 11 | 0.206 | 20.6 |
| 12 | 0.213 | 21.3 |
| 13 | 0.265 | 26.5 |
| 14 | 0.132 | 13.2 |
